# Genetic contribution to asthma informs acute chest syndrome pathophysiology and risk stratification

**DOI:** 10.1101/2025.09.30.25336662

**Authors:** Sara El Aouhel, Vanessa Bellegarde, Stennio Da Silva Faria, Tristan St-Laurent, Estelle Lecluze, Anne-Laure Pham Hung d’Alexandry d’Orengiani, Frédéric Galactéros, Pablo Bartolucci, Marc-André Legault, Guillaume Lettre, Thomas Pincez

**Author notes:** **Correspondence:** Thomas Pincez, Division of Hematology-Oncology, 3175 Chemin de la Côte Sainte-Catherine, CHU Sainte-Justine, Montreal, H3T 1C5 Quebec, Canada. **Declaration of Interest:** Thomas Pincez: Research funds from Biossil Inc. Pablo Bartolucci: - Consultant for ADDMEDICA, NOVARTIS, ROCHE, GBT, Bluebird, EMMAUS, HEMANEXT, AGIOS. - Lecture fees for NOVARTIS, ADDMEDICA, JAZZPHARMA. - Steering committee for NOVARTIS, ROCHE, ADDMEDICA, PFIZER - Research support from ADDMEDICA, foundation Fabre, NOVARTIS, Bluebird, EMMAUS - Cofounder and CSO of INNOVH The other authors have no potential conflicts to disclose.

## Abstract

Acute chest syndrome (ACS) is a severe complication of sickle cell disease (SCD), but we lack tool to identify patients at high risk of ACS. Epidemiological studies have found an association between asthma and ACS but whether this link is causal is unclear. We used polygenic score (PGS) to analyze whether the genetic susceptibility to asthma was associated with ACS and could be used to stratify the risk of ACS. We identified that a PGS for asthma (PGS_asthma_) was associated with ACS rate, but not ACS occurrence, in both the CSSCD and the GEN-MOD cohorts, independently of fetal hemoglobin (HbF) (β=0.17, standard error=0.06, p=0.006). This effect was mainly found in patients with HbF <5%. Combining PGS_asthma_ and HbF allowed to identify a population at high risk of ACS recurrence: individuals within the highest PGS_asthma_ quintile and the lowest HbF quintile. Partitioned PGS suggested that lymphocytes were the main driver of the genetically mediated risk of ACS by asthma. Finally, we assessed asthma and ACS overall genetic correlation. We found that these two conditions only partially overlap distinct, suggesting that asthma is not the main determinant of genetic propensity to ACS. In sum, our result suggests that patients with high genetic propensity to asthma are prone to recurrent ACS if not protected by high HbF levels. Combining PGS_asthma_ and HbF may allow identifying high risk patients for personalize ACS management. Apart from this population at high risk of ACS, additional genetic determinants independent from asthma contribute to ACS.

## Introduction

Sickle cell disease (SCD) is one of the most frequent monogenic diseases worldwide. SCD is due to a variant in the β-globin gene leading to an abnormal hemoglobin –hemoglobin S– which polymerizes while deoxygenated.^1^ The resulting hemolysis and vaso-occlusion cause several complications, including pain crisis, stroke, early organ damage, and acute chest syndrome (ACS).^2^ SCD is highly heterogeneous in terms of clinical severity and there is currently no tool to stratify the patients according to the risk of complications.^3^

ACS is a frequent complication in SCD, especially in young children, but can occur throughout life.^3^ Despite great improvement in management in the last decade, ACS remains potentially life-threatening, particularly in adults.^4,5^ Some patients will experience frequent ACS but the cause for these recurrent events is unclear.^42^ ACS is a heterogeneous syndrome defined as the occurrence of a new pulmonary consolidation associated with respiratory symptoms.^6^ Various non-mutually exclusive triggers can lead to ACS, including infection, vaso-occlusive pain crisis, fat embolism, and asthma.^7^ Several studies identified an epidemiological link between asthma and ACS. Children and adults with asthma have an increased risk of ACS.^8–12^ Asthma is considered as a frequent trigger of ACS in children.^13^ However, the causality of the association and the pathophysiological link between these conditions are unclear.

Genetic susceptibility to asthma has been extensively investigated by genome-wide association studies (GWAS), identifying many loci associated with asthma risk.^14–16^ The genetic susceptibility to ACS is much less known and the few loci found have not been replicated.^17,18^ Whether the genetic susceptibility of asthma and ACS is similar is unknown. Moreover, polygenic scores (PGS) summarizing the genetically determined susceptibility to asthma allowed to stratify the risk of asthma.^14^ Whether asthma PGS may allow identifying patients with SCD at high risk of ACS –allowing tailoring management accordingly– has not been investigated.

Here, we leveraged large asthma GWAS from non-SCD individuals and genotyped cohorts of children and adults with SCD to investigate the link between asthma and SCD. We hypothesized that the knowledge gained on asthma genetics can be used to improve our understanding and risk modeling of ACS. We aimed to assess whether asthma was causally associated with ACS and which pathophysiological component was involved. We also aimed to assess the utility of PGS for asthma (PGS_asthma_) in the risk stratification of ACS in patients with SCD.

## Methods

### Populations

We used the main genotyped dataset from the Cooperative Study of Sickle Cell Disease (CSSCD) as the discovery cohort.^19^ The CSSCD was a US multicentric prospective study of the natural history of SCD. The main genotyped dataset included 1,278 patients (mean age at inclusion, 14 ± standard deviation [SD] 12 years) of whom 662 (52%) were female (**Table S1**). A total of 484 (38%) experienced an ACS. The mean ACS rate (defined as the number of ACS per year of study) was 0.13 ± 0.29. As replication cohorts, we used the adult Genetic Modifier (GEN-MOD, n=406) cohort and a smaller subset of the CSSCD that has been genotyped separately, herein called CSSCD2 (n=261). GEN-MOD is an ongoing prospective cohort of adults with SCD in Henri-Mondor hospital, France.^20,21^ No participant of the three cohorts received hydroxyurea. The studies were approved by the institutional ethics committee, and we collected data according to the Helsinki declaration. DNA genotyping and data processing have been described elsewhere.^21,22^

### PGS_asthma_

For this study, we selected a recently published PGS_asthma_ (n=3,972,232 variants, PGS catalog number PGS004725)^23^ built using PRSmix+ from 47,314 individuals (n=8,043 cases). PRSmix+ improves prediction accuracy by using published PGS and independent biobanks to increase sample size and trans-ancestry transposability.^24^ We used plink2 to compute PGSasthma for each cohort,^25^ and then we normalized the PGS_asthma_ and used the obtained z-scores throughout the study.

We computed the effect of the PGS_asthma_ on ACS rate using quasi-Poisson regression in a model including age, sex, fetal hemoglobin (HbF), and the 10 first principal components. We used quasi-Poisson to be consistent with the previous studies investigating ACS and pain rates.^26^ However, we also performed a sensitivity analysis using more conservative linear regression. For linear regression, we applied inverse normal transformation to normalize the ACS rate. We also tested the association between PGS_asthma_ and other complications using quasi-Poisson (pain rate) or logistic (stroke, osteonecrosis, priapism, leg ulcer, and death) regression with the same covariates.

### PGS partitioning

We used a recently published partitioned PGS (pPGS) for asthma to analyze the different pathophysiological mechanisms of asthma. The pPGS was built using cell epigenomic data (H3K4me2 marks determined by chromatin immunoprecipitation followed by DNA sequencing (ChIP-seq)) and identified five clusters with different associations with asthma endophenotypes.^27^ We tested the association between the five pPGS clusters and asthma as described for the PGS_asthma_.

### GWAS and single-nucleotide polymorphisms (SNPs)-specific analyses

We performed a GWAS for ACS rate on the CSSCD using plink2 after adjusting ACS rate for sex and age and applying inverse normal transformation.^25^ We tested an additive model using linear regression adjusted for the 10 first principal components. We also focused on the SNPs located within the 17q21 locus and associated with asthma in Open Targets and GWAS catalog.^23,28^ We performed the same process as the GWAS for these SNPs.

### Genetic Correlation

To compare the genetic susceptibility to asthma and ACS, we used the African-ancestry results of the Global Biobank Meta-analysis Initiative, which included 32,658 individuals (n=5,051 cases) from 6 biobanks.^14^

We used GPS to analyze the genetic correlation between asthma and ACS GWAS. GPS has been developed for rare diseases in which the classical methods based on linkage disequilibrium are not suitable (heritability of ACS in the CSSCD cohort: h^2^, −0.74; SE, 0.55).^29^ GPS is a nonparametric test of the independence of the p-values obtained by the two GWAS to compare. We used the gps_cpp package to compute GPS and generated a control distribution using 1,000 permutations to obtain p-value.^30^

We analyzed the distance between the statistically suggestive SNPs (<1×10^-6^) found in the two GWAS using bedtools.^31^ We computed the correlation between the -log_10_ of the p-values using Spearman correlation coefficient.

### Statistical Analyses

We compared continuous and categorical variables using non-parametric Fisher’s exact and Wilcoxon-Mann-Whitney tests, respectively. All tests were two-sided, and a statistical significance threshold was set to p <0.05. We corrected for multiple testing the analysis of the *ORMDL3* locus with the Benjamini-Hocheberg method, using a 0.05 false discovery rate, and considered a q-value <0.05 as statistically significant.^32^ All statistical analyses were performed using RStudio (version 1.2.5033) or GraphPad Prism (version 9.2.0, GraphPad Software, LLC, CA).

## Results

### Genetically determined asthma risk modulates the rate of ACS

We used a PGS_asthma_ built from a large multi-ancestry population.^24^ In the CSSCD cohort, PGS_asthma_ was associated with ACS rate, independently of HbF (β=0.17, standard error [SE]=0.06, p=0.006) (**Table 1**). Of note, the association was specific to ACS rate as we found no association with ACS occurrence in a logistic regression model (β=0.07, SE=0.06, p=0.24). We found no association of PGS_asthma_ with pain rates, stroke, osteonecrosis, priapism, leg ulcers, and death (p≥0.17, **Table S2**). We integrated HbF into our model, as this is the major modifier of SCD complications, including ACS risk. We noted that the association was stronger in a model with HbF than without (β=0.13, SE=0.06, p=0.02). This is evocative of a differential effect depending on HbF level, despite the interaction term between HbF and the PGS_asthma_ was not significant (p=0.28). We previously shown that the effect of a PGS for HbF depended on the HbF level and was higher in individuals with low HbF levels. We thus separated our cohort into subgroups with HbF < and ≥5% (n=513 and n=625 respectively). We found that the effect of PGS_asthma_ was higher in patients with HbF <5% than in the whole cohort (β=0.24, SE=0.08, p=0.005) whereas we found no association in patients with HbF ≥5% (β=0.08, SE=0.09, p=0.37).

**Table 1.**
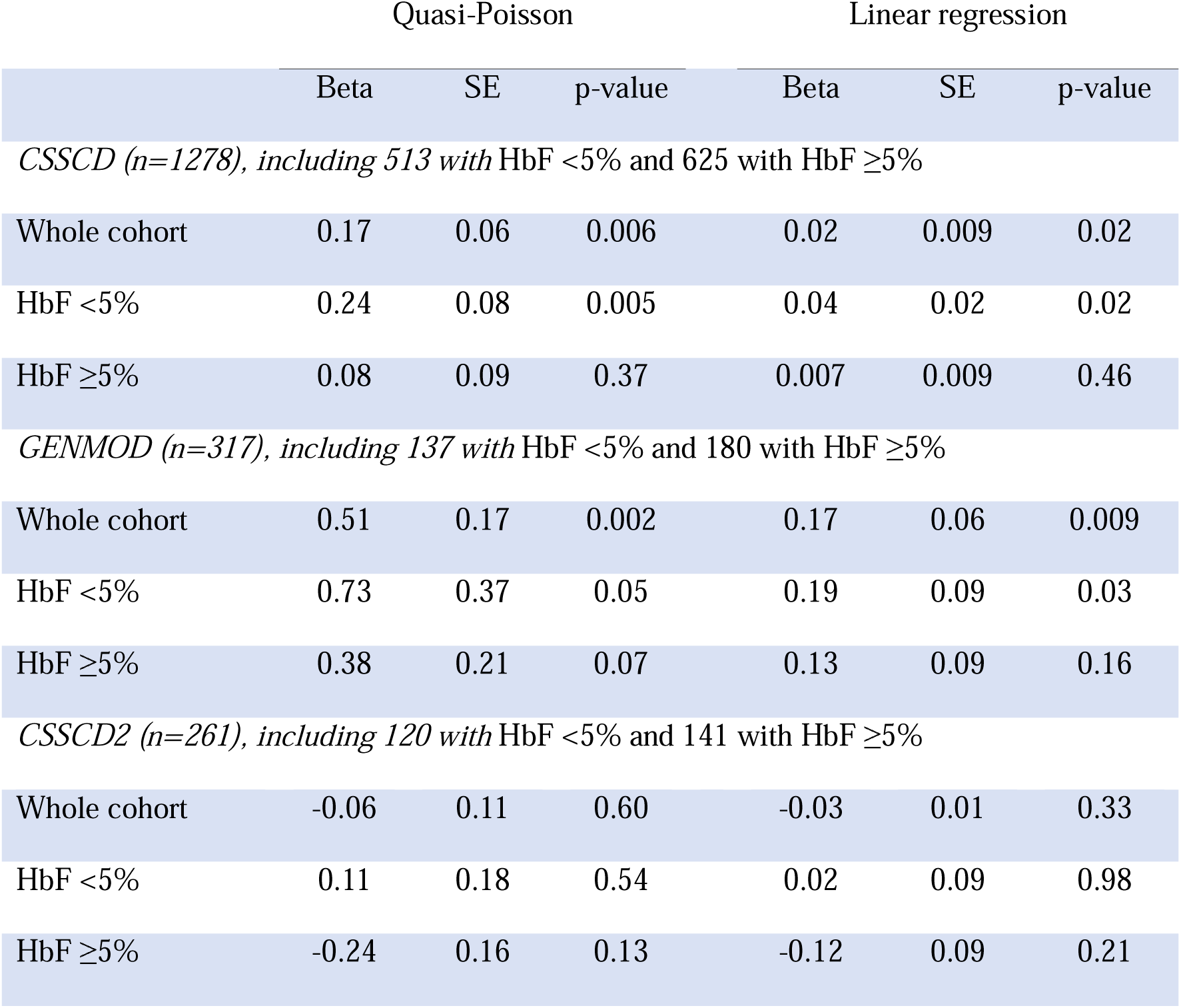
Association of the PGS_asthma_ with ACS rate. We tested the effect of PGS_asthma_ z-scores on ACS rate. We adjusted all models on age, sex, HbF and the 10 first principal component. We applied inverse normal transformation to normalize ACS rate in the linear regression models. HbF, fetal hemoglobin; SE, standard error.

We then aimed to validate our findings in other cohorts. In the adult GENMOD cohort (n=261), we found similar effects of the PGS_asthma_ on ACS rate (β=0.51, SE=0.17, p=0.003, adjusted on HbF). We also confirmed the association was stronger in patients with HbF <5% than in those with HbF>5% (β=0.73, SE=0.37, p=0.05 and β=0.38, SE=0.21, p=0.07, respectively). We found the same direction of effect in the smaller CSSD2 cohort for HbF <5% (n=120) but without significant association, possibly due to a lack of power.

In a sensitivity analysis, we used the more conservative linear regression approach on inverse transformation-normalized ACS rates. We found similar effects for all the models in each cohort (**Table 1**).

In sum, PGS_asthma_ is associated with ACS rate independently of HbF and this association is stronger in the subgroup of patients with baseline HbF <5%.

### PGS_asthma_ allows identifying ACS rate subgroups

We then asked whether combining PGS_asthma_ and HbF would allow stratifying the risk of ACS rate (**Figure 1A**). As PGS_asthma_ was associated with ACS rate and not occurrence, we analyzed the risk of ACS recurrence by studying the subgroup with at least one ACS. We separated the patients of the CSSCD based on PGS_asthma_ and HbF quintiles. We found that among patients within the lowest HbF quintile, those within the top PGS_asthma_ quintile had a higher ACS rate than those within the lowest PGS_asthma_ quintiles (median 0.35 vs. 0.23, p=0.049) (**Figure 1B**). In the lowest HbF and lowest PGS_asthma_ quintiles group, 2/21 patients (9.5%) had an ACS rate >0.5 compared to 10/26 (38%) in the lowest HbF and top PGS_asthma_ quintiles group (p=0.04) (**Figure 1C**). Within the highest HbF quintile, there was no difference in ACS rate between patients within the highest and the lowest PGS_asthma_ quintiles (median 0.24 vs. 0.27, p=0.24) (**Figure 1B**).

**Figure 1.**
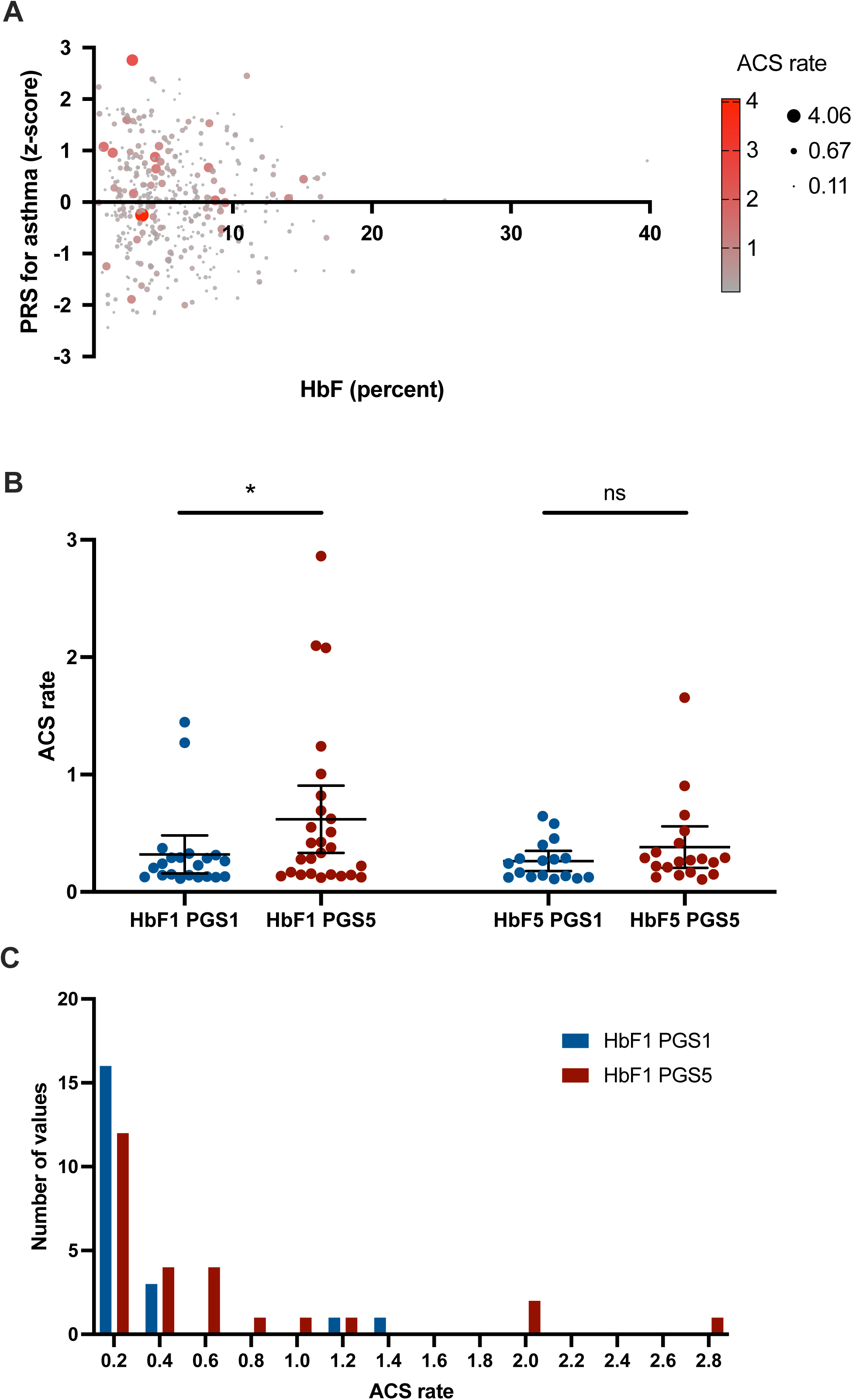
Combination of PRS for asthma and HbF identify a subgroup at high risk for asthma. A) ACS rate according to z-score of PGS_asthma_ and HbF value. Higher ACS rates are shown in red and larger circle. B) ACS rate according to HbF quintile and PGS_asthma_ quintile (1 being the lowest and 5 the highest quintile). C) Frequency of ACS rate value in the subgroup of patients within low HbF quintile within high or low PGS_asthma_ quintile. ACS, acute chest syndrome, HbF, fetal hemoglobin; PRS, polygenic risk score.

In sum, combining HbF and PGS_asthma_ allowed to identify a subgroup of patients at high risk of ACS recurrence.

### Causal role of asthma on ACS could be mediated by lymphocytes

To gain insight into the pathophysiological mechanisms underlying the effect of asthma on ACS, we used pPGS which was built by assigning asthma GWAS loci to the likely affected cell types based on the intersection between the GWAS loci and epigenomic H3K4me2 marks obtained by ChIP-seq.^27^ The pPGS identified five clusters with enrichment in CD4+ T-helper lymphocytes (cluster 1), neutrophils, eosinophils and mast cells (cluster 2), CD8+ T cells and B cells (cluster 3), lung epithelial cells (cluster 4) mast cells and activated group 2 innate lymphoid cells (cluster 5).

In the CSSCD, we found that the pPGS for clusters 1 and 3, the two clusters that included lymphocytes, were associated with ACS rate (**Table 2**). The effects of the two clusters were independent, as confirmed in a multivariate model including the five pPGS clusters. We found the same results for pPGS in the CSSCD using linear regression (**Table S3**). However, we did not replicate our findings in the GEN-MOD cohort (Cluster 1: Beta 0.17, SE 0.16, p=0.30, Cluster 3: Beta −0.04, SE 0.16, p=0.80).

**Table 2.** Association of the different cluster of the pPGS for asthma with ACS rate. We tested the effect of the clusters of the pPGS for asthma z-scores on ACS rate in the CSSCD using quasi-Poisson regression. We adjusted all models on age, sex, HbF and the 10 first principal component. We tested each cluster independently (left) and all together (right). HbF, fetal hemoglobin; SE, standard error.

|  | Single-cluster analysis |  |  | All-clusters analysis |  |  |
| --- | --- | --- | --- | --- | --- | --- |
|  | Beta | SE | p-value | Beta | SE | p-value |
| Cluster 1 | 23.35 | 9.79 | 0.02 | 0.013 | 0.06 | 0.02 |
| Cluster 2 | -0.01 | 0.06 | 0.84 | -0.01 | 0.06 | 0.82 |
| Cluster 3 | 0.13 | 0.06 | 0.03 | 0.13 | 0.06 | 0.03 |
| Cluster 4 | 0.04 | 0.05 | 0.43 | 0.04 | 0.05 | 0.56 |
| Cluster 5 | -0.06 | 0.06 | 0.25 | -0.07 | 0.06 | 0.24 |

As cluster 3 includes the well-known asthma risk locus 17q21 which contains the *ORMDL3* gene, we tested the association between SNPs at this locus and ACS rate and found no association (**Table S4**).

### Asthma and ACS are largely genetically distinct conditions

Having shown that PGS_asthma_ was causally associated with ACS, we finally investigated to what extend the genetic propensity to asthma and ACS were similar. We performed a GWAS for ACS rate in the CSSCD (**Figure S1**).^14^ To limit ancestry-related bias, we used the African-ancestry GWAS of the Global Biobank Meta-analysis Initiative for comparison with the ACS rate GWAS.^14^

We first compared the overall genetic correlation using GPS and found no correlation between the two GWAS (GPS=1.97, p=0.77) (**Figure 2A**).

**Figure 2.**
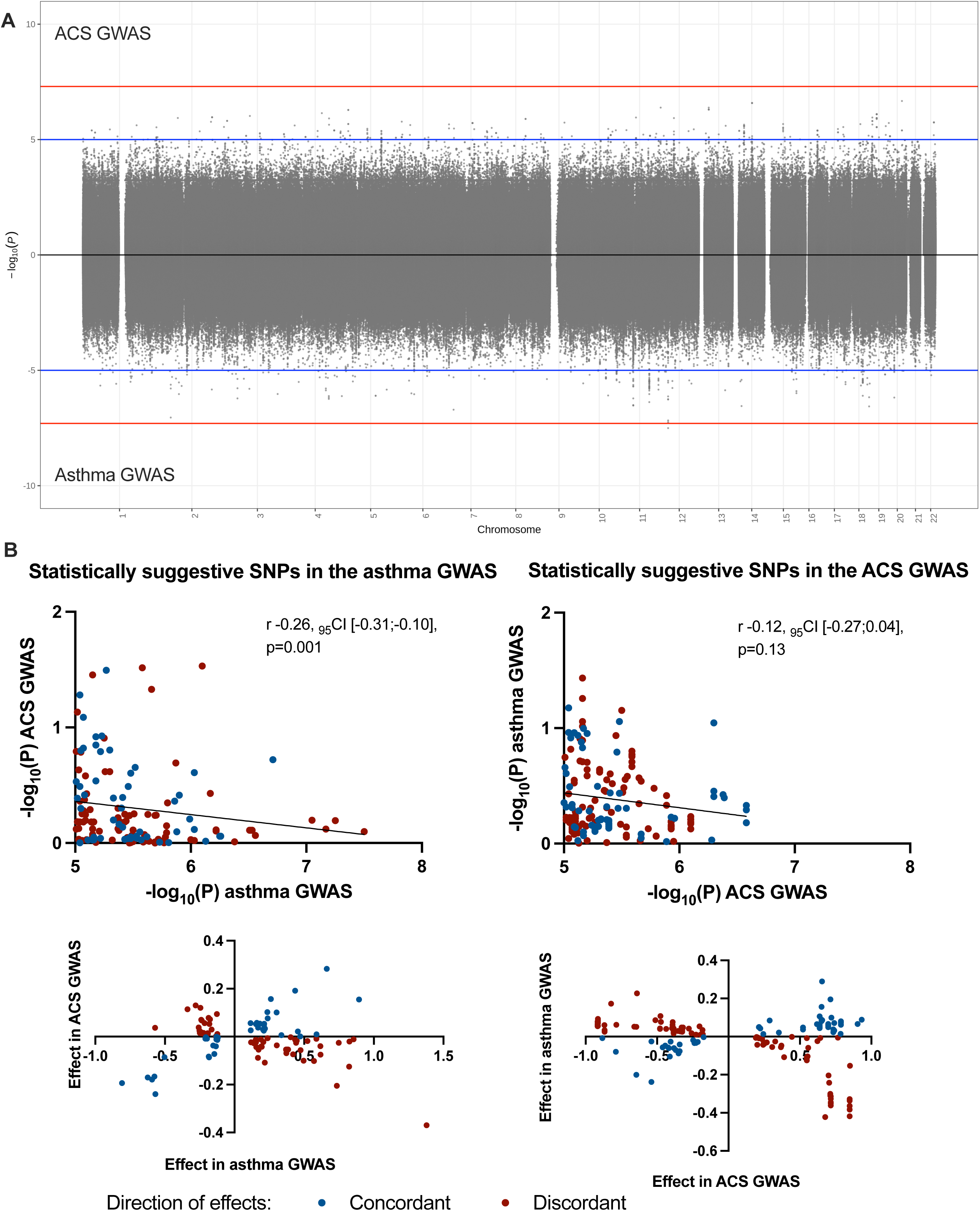
Genetic correlation between asthma and ACS. A) Miami plot of the GWAS for asthma and ACS. B) Comparison of p-values (upper panel) or effect size (lower panel) between asthma and ACS GWAS considering the SNPs statistically suggestive association (<1×10^-6^) in the asthma (left panel) or ACS (right panel) GWAS. The black line in the upper panel represents the linear regression line. ACS, acute chest syndrome.

We then focused on the statistically suggestive SNPs (p<1×10^-6^) from the two GWAS (n=124 in the ACS GWAS and n=149 in the asthma GWAS). We found that the closest SNPs were separated by 3.0 mega base pairs, thus in distinct loci unlikely to be in linkage disequilibrium. We found no positive correlation between the two GWAS. In fact, the p-values were inversely correlated when considering the statistically suggestive SNPs from the asthma GWAS (r −0.26, 95% confidence interval [CI] −0.31 to −0.10, p=0.001) (**Figure 2B**). The direction of the effects was discordant between the GWAS with a significant depletion for both GWAS (considering the statistically suggestive SNPs from the asthma GWAS: 37% with concordant effect, p=8.6×10^-4^; considering those from the ACS GWAS: 36% with concordant effect, p=0.001).

Thus, there is no overall genetic correlation between ACS and asthma, and the most significant loci are different. This suggests that although captured by PGS_asthma_, ACS and asthma genetics are only partially shared: some genetic determinants are specific to asthma and other to ACS.

## Discussion

Using PGS, we showed that asthma was causally associated with ACS rate, independently of the major genetic modifier HbF. This effect was mainly identified in patients with HbF <5%. The combination of HbF and PGS_asthma_ allowed identifying a subgroup of patients at high risk of ACS recurrence. Epigenomic-based pPGS suggested that the lymphocyte-mediated contribution to asthma was the main driver of ACS risk. Finally, we found that genetic propensity to asthma and ACS were only partially overlapping with genetic determinants unique to each condition.

ACS is driven by vaso-occlusion within the pulmonary microvasculature.^33^ This vaso-occlusion leads to hypoxia, endothelial injury, vasoconstriction, and eventually pulmonary parenchyma infarction.^33,34^ As a result, local acidosis and inflammation develop. All these consequences of vaso-occlusion will tend to further increase vaso-occlusion. This vicious cycle explains the tendency of ACS to spontaneously evolve toward a worsening. Clinical deterioration can be rapid in patients and ACS is a significant cause of death in SCD.^35,36^ Multiple triggers can contribute to pulmonary vaso-occlusion.^33^ In the patients presenting initially with vaso-occlusive pain episode, fat embolism or micro thrombi from the site of initial vaso-occlusion can lead to the secondary occurrence of ACS. Infection is another common trigger, especially in children, and both viral and bacterial agents can be involved.

The role of asthma as a trigger is supported by many epidemiological studies that identified a higher risk for ACS in children but also adults.^8–12^ Most studies also identified a higher risk of vaso-occlusive pain episodes.^12,37,38^ As ACS and vaso-occlusive pain episodes are risk factor for mortality in SCD,^35,39^ asthma has also been associated with an increased mortality risk.^40,41^ Our data show that, in addition to being a marker of severity, asthma is causally associated with the repetition of ACS. The fact that we found an association of PGS_asthma_ only with ACS rates and not occurrence suggests that the most striking consequence of asthma is SCD is the repetition of ACS. Indeed, around half of patients with SCD will present at least one ACS during their lifetime, but only a minority will present several episodes.^3^ In the CSSCD, among the 538 patients with ACS, 443 (82%) experienced only one episode.^42^ Thus, most ACS are likely caused by a transient trigger, whereas patients with asthma are prone to ACS all their lifetime and will develop several episodes.

The association of PGS_asthma_ with ACS rates but not occurrence also provides meaningful insight into an unexplained observation in ACS. Several multicenter studies identified that children hospitalized for an ACS before the age of 4 had a distinct lung evolution than those hospitalized for an ACS at an older age.^10,41^ Early onset ACS was associated with a high risk of ACS recurrence, asthma, and hospitalization for vaso-occlusive pain or ACS. DeBaun and Strunk hypothesized that early onset ACS resulted in an asthma-like syndrome, as can occur after viral infection at a young age.^6^ This asthma-like syndrome would further predispose regional hypoxia and inflammation upon infection or other airway triggers. Our data support this hypothesis by showing that a genetically determined risk of asthma results in a high risk of recurrence. This suggests that patients with a high propensity to asthma are more likely to develop hypoxia and inflammation upon common airway triggers.

Asthma pathophysiology is complex and heterogeneous across individuals.^43^ It associates atopy, bronchial hyperresponsiveness, and airway inflammation and remodeling.^44^ The various components involved drive different endotypes with specificity in terms of pathophysiological mechanisms. The Global Initiative for Asthma defines Type 2 inflammation phenotype characterized by interleukin (IL) 4, IL 5, IL 13 and eosinophils, and non-type 2 inflammation characterized by neutrophils.^45^ Asthma GWAS identified many loci and involving several immune-related pathways, such as IL 33 and IL1R1.^46^ Epigenomic-derived pPGS showed that this genetic propensity to asthma affected different immune cell types and was associated with distinct phenotypes.^27^ Our data suggest that lymphocytes could be the main cell types involved in the increased rate of ACS associated with asthma. These results warrant confirmation, as we could not replicate our findings in the GEN-MOD cohort, possibly due to the smaller sample size. However, they are consistent with the known pathophysiology of ACS, in which lymphocyte-associated inflammation is known to play a role, whereas the role of eosinophils is not established.^33,47^

We showed that genetically determined propensity to asthma increased ACS rate. However, genetic correlation analyses suggest that genetic determinants of ACS and asthma are only partially overlapping. The sample size of our SCD cohort was sufficient to use the GPS approach,^29,30^ allowing the results to be considered as having a low risk of false negatives. This result suggests that asthma is not the main mechanism predisposing to ACS. Patients with a high propensity to asthma are prone to develop multiple ACS but several other mechanisms and triggers can lead to ACS in patients with a low propensity to asthma. The lack of correlation and replication of the top asthma hits also suggest that not all the mechanisms underlying asthma are involved in ACS, which is consistent with our results with pPGS. Asthma is a heterogeneous disease, and the underlying pathophysiology may differ across patients.^43,44^ Our results suggest that not all the genetically mediated risk of asthma may increase the risk of ACS. Conversely, a genetic component may increase ACS risk through a different mechanism than asthma.

We found that the association of PGS_asthma_ with ACS was mainly in patients with HbF <5%. This suggests that patients with higher HbF levels may be protected from the increased risk of ACS conferred by genetic propensity to asthma. HbF is known to be the major modifier of SCD.^48^ Higher HbF levels have been associated with a reduced risk of most SCD complications and mortality.^35,39,42,49^ However, how HbF interacts with other risk factors remains poorly known. Our result suggests that the conequences on hypoxia and inflammation that can occur in patients with a high propensity to asthma depends on HbF levels.

The high heterogeneity of SCD is poorly understood and a major clinical challenge.^1^ Some patients rarely experience any complications, while others have a lot of them, as mentioned earlier for ACS.^42^ Thus, risk stratification is a major need in SCD care to identify patients requiring specific management or therapy intensification. Treatment options are expanding for SCD, but their role and the patients who would benefit from them are not yet clear.^50^ Risk stratification also allows to select patients at higher risk of complications for inclusion in clinical trials, an area of active research. With the notable exception of transcranial Doppler to stratify the risk of stroke,^51,52^ no tools currently exist in clinical practice. Despite its association with most complications, HbF is not effective for risk stratification, as most patients have low levels of HbF,^48^ impeding the identification of a high-risk population. Moreover, the cellular distribution of HbF among erythrocytes varies among patients, limiting the individual correlation between HbF level and SCD severity.^48,53^ PGS have been shown to be effective at identifying individuals at high risk of unfavorable outcomes in many settings, including common conditions such as diabetes but also rare conditions such as telomere biology disorders.^54,55^ In SCD, we have previously shown that a PGS for HbF improved risk modeling for pain rates.^56^ However, the clinical usefulness of PGS in ACS remains to be clarified. Here, we showed that the combination of HbF and PGS_asthma_ allowed to identify a subpopulation at high risk of ACS recurrence after a first episode. Given the potential severity of ACS, this population is at high risk of significant morbidity and mortality. Our results suggest that after a first episode, PGS_asthma_ could be used to propose treatment intensification in selected patients. While SCD-specific treatments should be considered, asthma-directed therapies may also warrant considerations. Further studies should investigate the optimal strategy for this high-risk population.

In conclusion, our data clarify the link between asthma and ACS. The genetic propensity to asthma has a causal effect on ACS rate in SCD. However, some genetic determinants are specific to asthma and others to ACS. This subset of shared genetic propensity is possibly mediated by lymphocytes, and affects both children and adults. As a result, patients with high genetic propensity for asthma are at higher risk of recurrent ACS if not protected by high HbF levels. The combination of HbF and PGS_asthma_ allows identifying a subpopulation that would benefit from therapy intensification. This study paves the way for the use of PGS as a stratification tool in SCD.

## Data Availability

All data produced in the present study are available upon reasonable request to the authors
and signed transfer agreement

## Acknowledgment

We thank all participants who contributed data to this study. This work was funded by the Canadian Institutes of Health Research (PJT #186159) and the Canada Research Chair program (G. Lettre).

## References

1. Ware RE, de Montalembert M, Tshilolo L, Abboud MR. Sickle cell disease. Lancet. 2017;390(10091):311-323. doi:10.1016/S0140-6736(17)30193-9

2. Kato GJ, Piel FB, Reid CD, et al. Sickle cell disease. Nat Rev Dis Primer. 2018;4(1):1–22. doi:10.1038/nrdp.2018.10

3. Castro O, Brambilla DJ, Thorington B, et al. The acute chest syndrome in sickle cell disease: incidence and risk factors. The Cooperative Study of Sickle Cell Disease. Blood. 1994;84(2):643–649.

4. Chaturvedi S, Ghafuri DL, Glassberg J, Kassim AA, Rodeghier M, DeBaun MR. Rapidly progressive acute chest syndrome in individuals with sickle cell anemia: a distinct acute chest syndrome phenotype. Am J Hematol. 2016;91(12):1185–1190. doi:10.1002/ajh.24539

5. Nouraie M, Darbari DS, Rana S, et al. Tricuspid regurgitation velocity and other biomarkers of mortality in children, adolescents and young adults with sickle cell disease in the United States: The PUSH study. Am J Hematol. 2020;95(7):766–774. doi:10.1002/ajh.25799

6. DeBaun MR, Strunk RC. The intersection between asthma and acute chest syndrome in children with sickle-cell anaemia. The Lancet. 2016;387(10037):2545–2553. doi:10.1016/S0140-6736(16)00145-8

7. Vichinsky EP, Neumayr LD, Earles AN, et al. Causes and outcomes of the acute chest syndrome in sickle cell disease. National Acute Chest Syndrome Study Group. N Engl J Med. 2000;342(25):1855–1865. doi:10.1056/NEJM200006223422502

8. Boyd JH, Macklin EA, Strunk RC, DeBaun MR. Asthma is associated with acute chest syndrome and pain in children with sickle cell anemia. Blood. 2006;108(9):2923–2927. doi:10.1182/blood-2006-01-011072

9. Knight-Madden JM, Forrester TS, Lewis NA, Greenough A. Asthma in children with sickle cell disease and its association with acute chest syndrome. Thorax. 2005;60(3):206–210. doi:10.1136/thx.2004.029165

10. DeBaun MR, Rodeghier M, Cohen R, et al. Factors predicting future ACS episodes in children with sickle cell anemia. Am J Hematol. 2014;89(11):E212–217. doi:10.1002/ajh.23819

11. Bernaudin F, Strunk RC, Kamdem A, et al. Asthma is associated with acute chest syndrome, but not with an increased rate of hospitalization for pain among children in France with sickle cell anemia: a retrospective cohort study. Haematologica. 2008;93(12):1917–1918. doi:10.3324/haematol.13090

12. Knight-Madden JM, Barton-Gooden A, Weaver SR, Reid M, Greenough A. Mortality, asthma, smoking and acute chest syndrome in young adults with sickle cell disease. Lung. 2013;191(1):95–100. doi:10.1007/s00408-012-9435-3

13. Vichinsky EP, Styles LA, Colangelo LH, Wright EC, Castro O, Nickerson B. Acute chest syndrome in sickle cell disease: clinical presentation and course. Cooperative Study of Sickle Cell Disease. Blood. 1997;89(5):1787–1792.

14. Tsuo K, Zhou W, Wang Y, et al. Multi-ancestry meta-analysis of asthma identifies novel associations and highlights the value of increased power and diversity. Cell Genomics. 2022;2(12):100212. doi:10.1016/j.xgen.2022.100212

15. Kim KW, Ober C. Lessons Learned From GWAS of Asthma. Allergy Asthma Immunol Res. 2018;11(2):170–187. doi:10.4168/aair.2019.11.2.170

16. Han Y, Jia Q, Jahani PS, et al. Genome-wide analysis highlights contribution of immune system pathways to the genetic architecture of asthma. Nat Commun. 2020;11(1):1776. doi:10.1038/s41467-020-15649-3

17. Pincez T, Ashley-Koch AE, Lettre G, Telen MJ. Genetic Modifiers of Sickle Cell Disease. Hematol Oncol Clin North Am. 2022;36(6):1097–1124. doi:10.1016/j.hoc.2022.06.006

18. Galarneau G, Coady S, Garrett ME, et al. Gene-centric association study of acute chest syndrome and painful crisis in sickle cell disease patients. Blood. 2013;122(3):434–442. doi:10.1182/blood-2013-01-478776

19. Farber MD, Koshy M, Kinney TR. Cooperative Study of Sickle Cell Disease: Demographic and socioeconomic characteristics of patients and families with sickle cell disease. J Chronic Dis. 1985;38(6):495–505. doi:10.1016/0021-9681(85)90033-5

20. Bartolucci P, Brugnara C, Teixeira-Pinto A, et al. Erythrocyte density in sickle cell syndromes is associated with specific clinical manifestations and hemolysis. Blood. 2012;120(15):3136–3141. doi:10.1182/blood-2012-04-424184

21. Ilboudo Y, Bartolucci P, Rivera A, et al. Genome-wide association study of erythrocyte density in sickle cell disease patients. Blood Cells Mol Dis. 2017;65:60–65. doi:10.1016/j.bcmd.2017.05.005

22. Bae HT, Baldwin CT, Sebastiani P, et al. Meta-analysis of 2040 sickle cell anemia patients: BCL11A and HBS1L-MYB are the major modifiers of HbF in African Americans. Blood. 2012;120(9):1961-1962. doi:10.1182/blood-2012-06-432849

23. Sollis E, Mosaku A, Abid A, et al. The NHGRI-EBI GWAS Catalog: knowledgebase and deposition resource. Nucleic Acids Res. 2023;51(D1):D977–D985. doi:10.1093/nar/gkac1010

24. Truong B, Hull LE, Ruan Y, et al. Integrative polygenic risk score improves the prediction accuracy of complex traits and diseases. Cell Genomics. 2024;4(4). doi:10.1016/j.xgen.2024.100523

25. Purcell S, Neale B, Todd-Brown K, et al. PLINK: A Tool Set for Whole-Genome Association and Population-Based Linkage Analyses. Am J Hum Genet. 2007;81(3):559–575. doi:10.1086/519795

26. Lettre G, Sankaran VG, Bezerra MAC, et al. DNA polymorphisms at the BCL11A, HBS1L-MYB, and beta-globin loci associate with fetal hemoglobin levels and pain crises in sickle cell disease. Proc Natl Acad Sci U S A. 2008;105(33):11869-11874. doi:10.1073/pnas.0804799105

27. Stikker B, Trap L, Sedaghati-Khayat B, et al. Epigenomic partitioning of a polygenic risk score for asthma reveals distinct genetically driven disease pathways. Eur Respir J. 2024;64(2):2302059. doi:10.1183/13993003.02059-2023

28. Ochoa D, Hercules A, Carmona M, et al. The next-generation Open Targets Platform: reimagined, redesigned, rebuilt. Nucleic Acids Res. 2023;51(D1):D1353–D1359. doi:10.1093/nar/gkac1046

29. Li YR, Li J, Zhao SD, et al. Meta-analysis of shared genetic architecture across ten pediatric autoimmune diseases. Nat Med. 2015;21(9):1018–1027. doi:10.1038/nm.3933

30. Willis TW, Wallace C. Accurate detection of shared genetic architecture from GWAS summary statistics in the small-sample context. PLOS Genet. 2023;19(8):e1010852. doi:10.1371/journal.pgen.1010852

31. Quinlan AR, Hall IM. BEDTools: a flexible suite of utilities for comparing genomic features. Bioinformatics. 2010;26(6):841–842. doi:10.1093/bioinformatics/btq033

32. Benjamini Y, Hochberg Y. Controlling the False Discovery Rate: A Practical and Powerful Approach to Multiple Testing. J R Stat Soc Ser B Methodol. 1995;57(1):289–300.

33. Gladwin MT, Vichinsky E. Pulmonary complications of sickle cell disease. N Engl J Med. 2008;359(21):2254–2265. doi:10.1056/NEJMra0804411

34. Gladwin MT, Rodgers GP. Pathogenesis and treatment of acute chest syndrome of sickle-cell anaemia. The Lancet. 2000;355(9214):1476-1478. doi:10.1016/S0140-6736(00)02157-7

35. Platt OS, Brambilla DJ, Rosse WF, et al. Mortality in sickle cell disease. Life expectancy and risk factors for early death. N Engl J Med. 1994;330(23):1639–1644. doi:10.1056/NEJM199406093302303

36. Darbari DS, Kple-Faget P, Kwagyan J, Rana S, Gordeuk VR, Castro O. Circumstances of death in adult sickle cell disease patients. Am J Hematol. 2006;81(11):858–863. doi:10.1002/ajh.20685

37. Cohen RT, Madadi A, Blinder MA, DeBaun MR, Strunk RC, Field JJ. Recurrent, severe wheezing is associated with morbidity and mortality in adults with sickle cell disease. Am J Hematol. 2011;86(9):756–761. doi:10.1002/ajh.22098

38. Glassberg JA, Chow A, Wisnivesky J, Hoffman R, Debaun MR, Richardson LD. Wheezing and asthma are independent risk factors for increased sickle cell disease morbidity. Br J Haematol. 2012;159(4):472–479. doi:10.1111/bjh.12049

39. Platt OS, Thorington BD, Brambilla DJ, et al. Pain in Sickle Cell Disease. N Engl J Med. 1991;325(1):11–16. doi:10.1056/NEJM199107043250103

40. Boyd JH, Macklin EA, Strunk RC, DeBaun MR. Asthma is associated with increased mortality in individuals with sickle cell anemia. Haematologica. 2007;92(8):1115–1118.

41. Vance LD, Rodeghier M, Cohen RT, et al. Increased risk of severe vaso-occlusive episodes after initial acute chest syndrome in children with sickle cell anemia less than 4 years old: Sleep and asthma cohort. Am J Hematol. 2015;90(5):371–375. doi:10.1002/ajh.23959

42. Vichinsky EP, Neumayr LD, Earles AN, et al. Causes and Outcomes of the Acute Chest Syndrome in Sickle Cell Disease. N Engl J Med. 2000;342(25):1855–1865. doi:10.1056/NEJM200006223422502

43. Kuruvilla ME, Lee FEH, Lee GB. Understanding Asthma Phenotypes, Endotypes, and Mechanisms of Disease. Clin Rev Allergy Immunol. 2019;56(2):219–233. doi:10.1007/s12016-018-8712-1

44. Holgate ST, Wenzel S, Postma DS, Weiss ST, Renz H, Sly PD. Asthma. Nat Rev Dis Primer. 2015;1(1):1–22. doi:10.1038/nrdp.2015.25

45. Global Initiative for Asthma. Global Strategy for Asthma Management and Prevention. Published online 2023.

46. Stikker BS, Hendriks RW, Stadhouders R. Decoding the genetic and epigenetic basis of asthma. Allergy. 2023;78(4):940–956. doi:10.1111/all.15666

47. Bhasin N, Sarode R. Acute Chest Syndrome in Sickle Cell Disease. Transfus Med Rev. 2023;37(3):150755. doi:10.1016/j.tmrv.2023.150755

48. Steinberg MH. Fetal hemoglobin in sickle cell anemia. Blood. 2020;136(21):2392–2400. doi:10.1182/blood.2020007645

49. Ohene-Frempong K, Weiner SJ, Sleeper LA, et al. Cerebrovascular accidents in sickle cell disease: rates and risk factors. Blood. 1998;91(1):288–294.

50. Salinas Cisneros G, Thein SL. Recent Advances in the Treatment of Sickle Cell Disease. Front Physiol. 2020;11. doi:10.3389/fphys.2020.00435

51. Adams RJ, McKie VC, Hsu L, et al. Prevention of a first stroke by transfusions in children with sickle cell anemia and abnormal results on transcranial Doppler ultrasonography. N Engl J Med. 1998;339(1):5–11. doi:10.1056/NEJM199807023390102

52. Verduzco LA, Nathan DG. Sickle cell disease and stroke. Blood. 2009;114(25):5117-5125. doi:10.1182/blood-2009-05-220921

53. Khandros E, Huang P, Peslak SA, et al. Understanding heterogeneity of fetal hemoglobin induction through comparative analysis of F and A erythroblasts. Blood. 2020;135(22):1957–1968. doi:10.1182/blood.2020005058

54. Ortega HI, Udler MS, Gloyn AL, Sharp SA. Diabetes mellitus polygenic risk scores: heterogeneity and clinical translation. Nat Rev Endocrinol. 2025;21(9):530–545. doi:10.1038/s41574-025-01132-w

55. Poeschla M, Arora UP, Walne A, et al. Polygenic modifiers impact penetrance and expressivity in telomere biology disorders. J Clin Invest. 2025;135(16). doi:10.1172/JCI191107

56. Pincez T, Lo KS, D’Orengiani ALPH d’Alexandry, et al. Variation and impact of polygenic hematological traits in monogenic sickle cell disease. Haematologica. 2023;108(3):870–881. doi:10.3324/haematol.2022.281180

