## Supplemental data for "Genetic contribution to asthma informs acute chest syndrome pathophysiology and risk stratification"

|  | CSSCD | GEN-MOD | CSSCD2 |
| --- | --- | --- | --- |
| Number of individuals | 1,278 | 317 | 261 |
| Sex, male/female | 616/662 | 141/176 | 120/141 |
| Age (year), mean ± SD | 14 ± 12 | 31 ± 9 | 17 ±11 |
| Pain rate, mean ± SD | 0.81 ± 1.42 | NA | NA |
| ACS rate, mean ± SD | 0.13 ± 0.29 | 0.17 ±0.48 | 0.09 ±0.18 |
| Stroke (%) | 105 (8.3) | NA | NA |
| Priapism (%) | 107 (8.3) | NA | NA |
| Leg ulcer (%) | 163 (12.7) | NA | NA |
| Death (%) | 44 (3.4) | NA | NA |
| HbF (%), mean ± SD | 6.5 ± 4.4 | 6.7 ±4.8 | 6.9 ±5.6 |
| Hydroxyurea use (%) | 0 (0) | 0 (0) | 0 (0) |

**Table S1. Characteristics of the cohorts included**

In the CSSCD cohort, painful episodes are considered when requiring emergency room visits. For pain and and acute chest syndrome (ACS), rates are defined as the number of episodes per year. NA, not available. SD, standard deviation

|  | Beta | SE | p-value |
| --- | --- | --- | --- |
| Pain rate | 0.02 | 0.09 | 0.81 |
| Stroke | 0.10 | 0.11 | 0.38 |
| Osteonecrosis | 0.007 | 0.08 | 0.93 |
| Priapism | 0.17 | 0.12 | 0.17 |
| Leg ulcer | -0.01 | 0.10 | 0.89 |
| Death | -0.05 | 0.17 | 0.78 |

**Table S2. Association of the polygenic risk score for asthma and complications**

We used quasi-Poisson (pain rate) or logistic regression (other variables) in models adjusted for age, sex, HbF and the 10 first principal components. We show the result for the normalized polygenic risk score.

|  | Beta | SE | p-value |
| --- | --- | --- | --- |
| Cluster 1 | 0.02 | 0.01 | 0.03 |
| Cluster 2 | -0.01 | 0.01 | 0.87 |
| Cluster 3 | 0.02 | 0.01 | 0.05 |
| Cluster 4 | 0.01 | 0.01 | 0.50 |
| Cluster 5 | -0.01 | 0.01 | 0.25 |

**Table S3. Association of the different cluster of the pPGS for asthma with ACS rate**

We tested the effect of the clusters of the pPGS for asthma z-scores on ACS rate z-scores in the CSSCD using linear regression. We adjusted all models on age, sex, HbF and the 10 first principal component. We tested each cluster independently.

| **SNP** | **Beta** | **SE** | **p-value** | **q-value** |
| --- | --- | --- | --- | --- |
| 17_39754115_C_T | 0.02 | 0.08 | 0.80 | 0.86 |
| 17_39756124_C_T | -0.10 | 0.10 | 0.30 | 0.46 |
| 17_39766006_G_A | -0.16 | 0.11 | 0.14 | 0.46 |
| 17_39781794_C_T | -0.12 | 0.10 | 0.22 | 0.46 |
| 17_39867492_A_G | -0.20 | 0.10 | 0.04 | 0.46 |
| 17_39869916_C_T | 0.09 | 0.08 | 0.26 | 0.46 |
| 17_39883866_T_C | -0.17 | 0.12 | 0.16 | 0.46 |
| 17_39887090_G_A | 0.09 | 0.08 | 0.24 | 0.46 |
| 17_39899863_T_C | -0.10 | 0.09 | 0.25 | 0.46 |
| 17_39900936_G_A | -0.17 | 0.12 | 0.14 | 0.46 |
| 17_39900944_G_A | 0.07 | 0.08 | 0.37 | 0.54 |
| 17_39905186_T_C | -0.20 | 0.12 | 0.08 | 0.46 |
| 17_39905943_G_A | -0.20 | 0.12 | 0.08 | 0.46 |
| 17_39905964_C_T | -0.14 | 0.10 | 0.17 | 0.46 |
| 17_39908152_CGG_C | -11.54 | 549.30 | 0.98 | 0.87 |
| 17_39908152_C_T | -0.17 | 0.12 | 0.14 | 0.46 |
| 17_39909987_T_C | 0.01 | 0.08 | 0.89 | 0.86 |
| 17_39910014_G_T | 0.10 | 0.08 | 0.23 | 0.46 |
| 17_39910119_T_C | 0.01 | 0.08 | 0.93 | 0.86 |
| 17_39911790_T_C | -0.04 | 0.10 | 0.66 | 0.79 |
| 17_39912261_T_C | 0.06 | 0.08 | 0.43 | 0.59 |
| 17_39913696_C_T | -0.06 | 0.09 | 0.55 | 0.72 |
| 17_39917793_G_A | -0.03 | 0.08 | 0.73 | 0.83 |
| 17_39919173_G_A | 0.01 | 0.08 | 0.86 | 0.86 |
| 17_39919884_T_G | 0.01 | 0.08 | 0.90 | 0.86 |
| 17_39924612_A_G | -0.12 | 0.08 | 0.13 | 0.46 |
| 17_39924659_G_A | -0.11 | 0.10 | 0.29 | 0.46 |
| 17_39933091_A_G | 0.04 | 0.08 | 0.60 | 0.75 |

**Table S4. Association of the 17q21 locus with ACS**

We adjusted ACS rate for sex and age and applying inverse normal transformation. We then tested an additive model using linear regression adjusted for the 10 first principal components. SE, standard error; SNP, single nucleotide polymorphism.

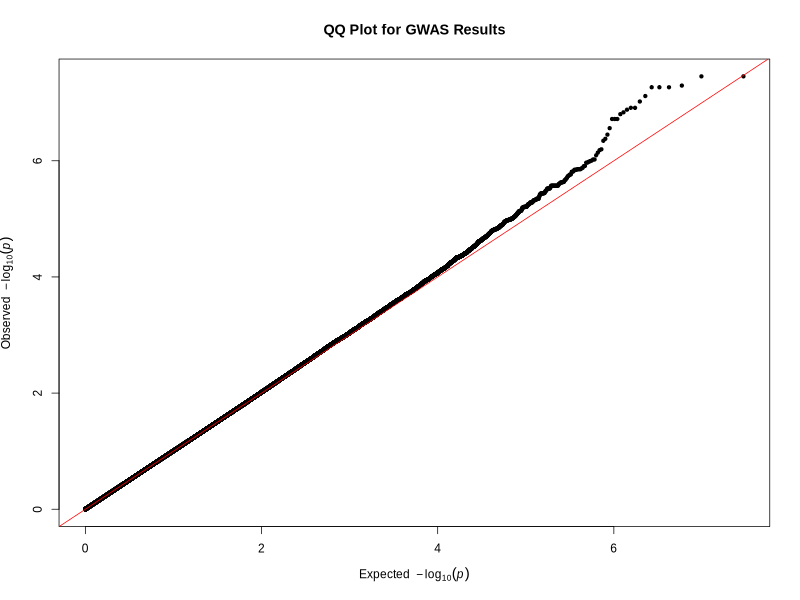

**Figure S1. Quantile-quantile plot for the GWAS of ACS rate in the CSSCD.** We used a linear regression model on normalized ACS rate, adjusted for age, sex, and the first 10 principal components. Lambda gc=1.011.
